# Automated detection of large vessel occlusion using deep learning: a pivotal multicenter clinical trial and reader assessment study

**DOI:** 10.1101/2024.04.24.24306331

**Authors:** Jae Guk Kim, Sue Young Ha, You-Ri Kang, Hotak Hong, Dongmin Kim, Myungjae Lee, Leonard Sunwoo, Wi-Sun Ryu, Joon-Tae Kim

## Abstract

**Background:** This multicenter clinical trial evaluated the stand-alone efficacy and the improvements in diagnostic accuracy of early-career physicians using a deep learning-based software to detect large vessel occlusion (LVO) in CT angiography (CTA).

**Methods:** This multicenter pivotal clinical trial included 595 ischemic stroke patients from January 2018 to September 2023. Standard reference and LVO locations (intracranial internal carotid artery [ICA], M1, or M2) were determined by consensus among three expert vascular neurologists after reviewing CTA, MR imaging, and symptom data. The performance of the JLK-LVO software was evaluated against a standard reference, and its impact on the diagnostic accuracy of four residents involved in stroke care was assessed. Performance metrics included the area under the receiver operating characteristic curve (AUROC), sensitivity, specificity, positive predictive value (PPV), and negative predictive value (NPV).

**Results:** Among the 595 patients (mean age 68.5 ± 13.4 years, 56% male), 275 (46.2%) had LVO. The median time interval from the last known well moment to the CTA was 46 hours (IQR 11.8 to 64.4). For LVO detection, the software demonstrated a sensitivity of 86% and a specificity of 97%. For isolated M2 occlusions, it achieved a sensitivity of 69% and a specificity of 96%. The reader assessment study showed that reading with software assistance improved the sensitivity by 4.0% and AUROC by 2.4% (all p < 0.001) compared to readings without AI assistance.

**Conclusion:** The software demonstrated a high detection rate for proximal LVO and moderate sensitivity for isolated MCA-M2 occlusion. In addition, the software improved diagnostic accuracy of early-career physicians in detecting LVO.

## Introduction

Advancements in stroke imaging and procedural devices have expanded the window for endovascular therapy (EVT) in patients with large vessel occlusion (LVO).^1^ Recent randomized clinical trials have established a new standard of care for patients with LVO who arrive at the hospital within 6 to 24 hours of their last known well time.^2, 3^ The triage process for these clinical trials primarily relies on magnetic resonance (MR) perfusion or computed tomography (CT) perfusion to identify clinical or tissue mismatches.^2, 3^ However, the majority of primary stroke facilities worldwide lack widespread access to these advanced imaging techniques.^4^ Recent studies have emphasized using more accessible imaging methods like CT angiography (CTA). The CT for Late Endovascular Reperfusion (CLEAR) trial found comparable clinical outcomes between patients selected using non-contrast CT with CTA and those selected with CT perfusion or MR perfusion.^5^ Furthermore, a sub-study conducted by the HERMES collaboration (Highly Effective Reperfusion Evaluated in Multiple Endovascular Stroke Trials) has expanded on this notion within the early time frame (0-6 hours), showing that the rates of favorable functional outcomes were similar between patients who underwent CT perfusion and those who did not.^6^

Initially, 66% of EVT candidates were directed to centers incapable of performing EVT,^7^ despite having better chances of favorable outcomes at EVT-capable sites. Consequently, it is crucial for non-EVT-capable centers to reliably and promptly detect LVO at all times—24 hours a day, 7 days a week—facilitating the swift transfer of patients to EVT-capable facilities. However, the scarcity of vascular experts poses a challenge for many small hospitals. Even in EVT-capable centers, the ability to screen CTA for the presence of LVO could improve efficiency, staffing, and the time from patient arrival to the initiation of the procedure by facilitating the detection of LVOs.

This multicenter pivotal clinical trial aimed to demonstrate the efficacy of a fully automated, deep learning-based software (JLK-LVO) in detecting LVO in patients with acute ischemic stroke. The algorithm’s performance was evaluated against a standard reference determined by stroke experts. Furthermore, we examined whether the use of artificial intelligence (AI) software enhances the diagnostic accuracy of early-career physicians compared to their performance without AI assistance.

## Method

### Study population

From January 2018 to September 2023, we included patients with ischemic stroke who were admitted to Chonnam National University and Daejeon Eulji University Hospital within 7 days of symptom onset. Among 603 eligible patients, we excluded 8 due to poor image quality or severe metallic artifacts (n = 4), insufficient contrast filling (n = 1), and source image thickness > 2 mm (n = 3), leaving 595 patients for analysis (see Supplementary Fig. 1). The study protocol was approved by the institutional review board of each hospital. Written consent was waived due to the study’s anonymous and retrospective design.

### Definition of large vessel occlusion

In this study, LVO was defined as arterial occlusion involving the intracranial segment of the internal carotid artery (ICA), the M1 segment of the middle cerebral artery (MCA-M1), and the M2 segment of the MCA (MCA-M2). The intracranial ICA refers to the segment of the ICA extending from the petrous part to the bifurcation of the MCA and the anterior cerebral artery (ACA).^8^ The MCA-M1 segment is defined as the portion of the MCA from the MCA-ACA bifurcation to the MCA branching point. The MCA-M2 segment refers to the portion of the MCA ascending vertically along the Sylvian fissure from the MCA branching point.^8^ Occlusions at the intracranial ICA or MCA-M1 were categorized as intracranial LVO, while occlusions at MCA-M2 were categorized as distal MCA occlusion in our study. In cases where the MCA divided early, we employed a functional rather than a traditional definition: the artery segment closest to its origin was designated as the M1 segment, and branches further downstream were classified as M2 segments.^9^ To ascertain the presence of LVO, two experienced vascular neurologists, each with at least 5 years of experience, meticulously reviewed the CTA source images, maximum intensity projection (MIP) images, and 3-dimensional rendering images, in addition to patients’ MR imaging (MRI) scans and symptom data. In instances of labeling disagreement, a third reviewer made the final decision.

### Deep learning-based software

Source images of CTA with slice thickness between 0.5 – 2 mm were fed into the commercially available deep learning-based software (JLK-LVO, JLK Inc., Seoul, Korea). In brief, an automated algorithm selects slices from source images to construct MIP images. The vessel segmentation involves a 2D U-Net based on the Inception Module,^10^ trained to segment vessels in axial MIP images. A vessel occlusion detection algorithm follows, involving the combination of vessel masks into a compressed image for training an EfficientNetV2 model.^11^

### Study design

#### Stand-alone performance

LVO probability scores predicted by software were used to evaluate the standalone performance metrics of the software, including the area under the receiver operating characteristic curve (AUROC), sensitivity, specificity, positive predictive value (PPV), and negative predictive value (NPV).

#### Reader assessment study

A retrospective, crossover-designed trial was conducted to assess the efficacy of software in aiding diagnostic decisions for detecting LVO in CTA. The multi-reader study involved four residents specializing in the care of ischemic stroke from the fields of radiology, neurology, neurosurgery, and emergency medicine. Before the evaluation, the reviewers were divided into groups A and B. Group A was provided with original CTA images, including MIP and 3D rendering images, along with the software’s results (with AI assistance; Supplementary Figure 2). Conversely, Group B received only the original CTA images, including MIP and 3D rendering images (without AI assistance). The software presented segmented and merged vessel images with a heatmap and an LVO probability score. A custom image viewer (JLK Inc., Seoul, Republic of Korea) was utilized to evaluate the CTA and the software’s output. The reviewers were blinded to the standard reference confirmed by expert consensus. A second assessment was conducted 4 weeks after a washout period, during which subjects were shuffled and randomly allocated new study numbers. In this second assessment, Group A reassessed the CTA images without the aid of the software’s findings (without AI assistance), while Group B reassessed the CTA images with the software’s results (with AI assistance). Readers determined the presence of LVO and assigned a confidence score using a 5-point scale. The study design is illustrated in Supplementary Figure 3.

### Statistical analysis

Using the t-test or rank sum test for continuous variables, and the chi-square test for categorical variables as appropriate, we compared baseline characteristics stratified by the presence of LVO or by participating centers. To evaluate the accuracy of the software in diagnosing LVO, we computed the AUROC, as well as sensitivity, specificity, PPV, and NPV. A 1000-repeat bootstrap analysis was employed to calculate the 95% confidence intervals (CIs) for all parameters. The AUROC was used in combination with the DeLong method^12^ to compute the standard error (SE) of the AUROC. The cutoff for the LVO probability score used in the analysis was set at 0.5. We conducted an additional analysis to determine the optimal threshold that would yield the maximum Youden index (sensitivity + specificity - 1). Given that the software is primarily intended for screening LVO, we also computed specificity, PPV, and NPV at sensitivity levels of 0.90 and 0.95.

After dividing the subjects into isolated MCA-M2 occlusion and intracranial LVO groups, we reran the analysis for subgroup analysis. In this analysis, patients without LVO were included as the control group for both subgroups. Taking into account the EVT time window, we repeated the analysis after excluding patients whose onset to CTA time exceeded 24 hours.

The Obuchowski-Rockette method was used for analyzing multireader multicase (MRMC) studies, along with the MRMCaov library,^13^ for all analyses of diagnostic performance in this study. This method tested the null hypothesis that the average AUROC of the readers without AI assistance was equal to that with AI assistance. The Obuchowski-Rockette method accounts for the fact that, in an MRMC study, the same cases are evaluated by each reader. Consequently, the error terms are assumed to be equi-covariant among readers and cases, rather than independent. We also calculated sensitivity, specificity, accuracy, and there differences between with and without AI assistance. A P value < 0.05 was considered statistically significant. The analyses were conducted using R version 4.2.3, STATA software (version 16.0, TX, USA) and MedCalc (version 17.2, MedCalc Software, Ostend, Belgium, 2017).”

## Results

### Baseline characteristics

Among 595 patients, 275 (46.2%) were diagnosed with LVO. Specifically, 213 patients had intracranial LVO, and 62 patients exhibited isolated MCA-M2 occlusion. Details regarding the occlusion sites are presented in Supplementary Table 1. The mean age was 68.5 (SD 13.4) and 332 (56%) were male. The median time interval between the last well known to CTA was 46 hours (interquartile range 11.8 to 64.4). Patients with LVO were of advanced age, experienced shorter time intervals between the onset of symptoms and imaging and presented with less severe strokes compared to those without LVO (Table 1). Analysis based on the participating centers revealed that participants from Daejeon Eulji University Hospital experienced significantly shorter delays between the onset of symptoms and imaging, and were more likely to receive endovascular treatment, compared to those from Chonnam National University Hospital (Supplementary Table 2).

**Table 1.** Baseline characteristics stratified by presence of large vessel occlusion.

|  | No LVO (n = 320) | LVO (n = 275) | P value |
| --- | --- | --- | --- |
| Age | 66.4 ± 13.1 | 71.2 ± 13.3 | < 0.001 |
| Sex, men | 180 (56.3%) | 152 (55.3%) | 0.81 |
| Onset to admission, hr | 6.8 (2.5 – 25.8) | 2.7 (1.1 – 9.3) | < 0.001<sup>a</sup> |
| Onset to imaging, hr | 49.2 (18.2 – 68.2) | 41.7 (4.7 – 60.3) | < 0.001<sup>a</sup> |
| Initial NIHSS | 2 (1 – 4) | 11 (6 – 15) | < 0.001<sup>a</sup> |
| Previous stroke | 51 (15.9%) | 50 (18.5%) | 0.42 |
| Hypertension | 181 (56.6%) | 158 (58.3%) | 0.67 |
| Diabetes | 98 (30.6%) | 70 (25.8%) | 0.20 |
| Atrial fibrillation | 53 (16.6%) | 111 (41.0%) | < 0.001 |
| Revascularization therapy | 40 (12.5%) | 120 (43.6%) | < 0.001 |
| Intravenous only | 27 (8.4%) | 22 (8.1%) |  |
| Endovascular only | 8 (2.5%) | 43 (15.9%) |  |
| Combined | 5 (1.6%) | 55 (20.3%) |  |

Data were presented as mean±SD, median (interquartile range), and number (percentage).
<sup>a</sup>Rank-sum test was used.
LVO=large vessel occlusion; NIHSS=National Institute of Health Stroke Scale.

### Stand-alone performance of software

The software achieved an AUROC of 0.961 (95% CI, 0.945 – 0.976; Figure 1A) at a cutoff point of 0.50. The sensitivity, specificity, PPV, and NPV were 0.858 (95% CI, 0.811 – 0.897), 0.969 (95% CI, 0.943 – 0.985), 0.959 (95% CI, 0.927 – 0.980), and 0.888 (95% CI, 0.850 – 0.919), respectively, as detailed in Table 2. The highest Youden index was observed at the optimal cutoff point of 0.362, with corresponding values of 0.880 (95% CI, 0.836 – 0.916) for sensitivity, 0.953 (95% CI, 0.924 – 0.974) for specificity, 0.942 (95% CI, 0.906 – 0.967) for PPV, and 0.902 (95% CI, 0.866 – 0.932) for NPV, respectively. At a given sensitivity of 0.90, the specificity was 0.916 (95% CI, 0.825 – 0.968).

**Figure 1.**
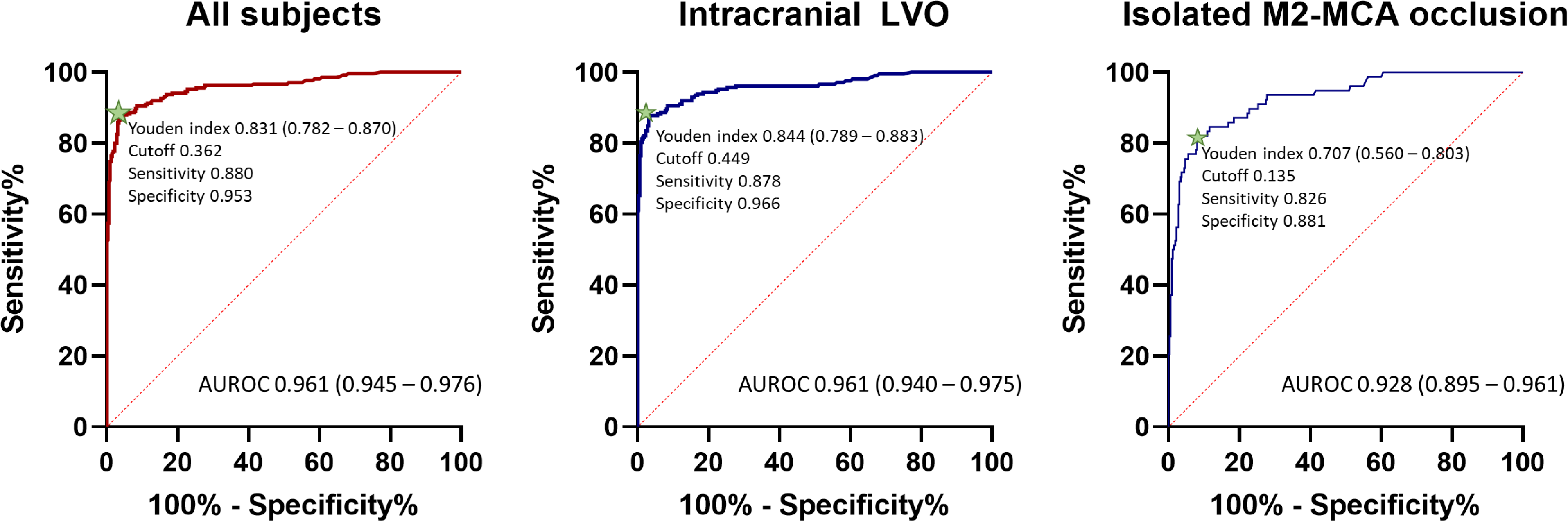
Diagnostic performance of software in all subjects and subgroup categorized into intracranial large vessel occlusion and isolated M2-MCA occlusion. Green starts indicate the cutoff point exhibiting the highest Youden index. MCA=middle cerebral artery; LVO=large vessel occlusion

**Table 2.** Diagnostic performance of software detecting large vessel occlusion.

|  | Confusion matrix | Prediction |  |
| --- | --- | --- | --- |
|  |  | LVO | No LVO |
| Threshold of 0.50 | Ground truth, LVO | 236 | 39 |
|  | Ground truth, no LVO | 10 | 310 |
|  | Sensitivity (95% CI) | 0.858 (0.811 – 0.897) |  |
|  | Specificity (95% CI) | 0.969 (0.943 – 0.985) |  |
|  | PPV (95% CI) | 0.959 (0.927 – 0.980) |  |
|  | NPV (95% CI) | 0.888 (0.850 – 0.919) |  |
|  | Youden (J) index (95% CI) | 0.833 (0.784 – 0.869) |  |
|  | J<sub>max</sub> cutoff point | 0.362 |  |
| Optimal threshold | J<sub>max</sub> Sensitivity (95% CI) | 0.880 (0.836 – 0.916) |  |
|  | J<sub>max</sub> Specificity (95% CI) | 0.953 (0.924 – 0.974) |  |
|  | J<sub>max</sub> PPV (95% CI) | 0.942 (0.906 – 0.967) |  |
|  | J<sub>max</sub> NPV (95% CI) | 0.902 (0.866 – 0.932) |  |
| Fixed sensitivity of 0.90 | Sens<sub>90</sub> Specificity (95% CI) | 0.916 (0.825 – 0.968) |  |
|  | Sens<sub>90</sub> PPV (95% CI) | 0.902 (0.860 – 0.934) |  |
|  | Sens<sub>90</sub> NPV (95% CI) | 0.916 (0.880 – 0.944) |  |
|  | Sens<sub>90</sub> cutoff point | 0.221 |  |

J<sub>max</sub> represents, across all thresholds, the maximum Youden index (sensitivity + specificity – 1). As a secondary reference point, J<sub>max</sub> provides an optimality criterion with equal weighting for sensitivity and specificity. LVO=large vessel occlusion; CI=confidence interval; PPV=positive predictive value; NPV=negative predictive value.

When limiting the analysis to intracranial LVO, the AUROC was 0.961 (95% CI: 0.940–0.975; Figure 1B). The sensitivity and specificity at the cutoff point of 0.5 were 0.873 (95% CI: 0.821 – 0.915) and 0.969 (95% CI: 0.943 – 0.985), respectively (Supplementary Table 3). Restricting the analysis to isolated MCA-M2 occlusion, the software demonstrated an AUROC of 0.928 (95% CI, 0.895 – 0.961; see Figure 1C), with a sensitivity of 0.692 and a specificity of 0.960 (95% CI, 0.932 – 0.978) and an NPV of 0.928 (95% CI, 0.895 – 0.954).

Considering the EVT time window, the analysis was restricted to those whose symptom onset to imaging was within 24 hours (n = 195), of which 109 (55.9%) demonstrated LVO. The software exhibited an AUROC of 0.973 (95% CI, 0.939 - 0.991; Supplementary Figure 4). With the cutoff point set at 0.5, the sensitivity and specificity were 0.890 (95% CI, 0.817 – 0.936) and 0.965 (95% CI, 0.902 – 0.991), respectively.

### Reader assessment study

In all reviewers, the sensitivities of reading with AI assistance versus without were 91.82% (95% CI, 90.04 – 93.37) and 87.81% (95% CI, 85.73 – 89.68), respectively, with a mean difference of 4.00% (95% CI, 2.17 to 5.84; p < 0.001; Table 3). The specificities were 95.70% and 96.30%, respectively, with no statistical difference observed. Reading with AI assistance yielded higher accuracy, with a mean difference of 0.76% (95% CI, 0.01 to 1.50; p = 0.049). The average AUROC with AI assistance was significantly higher compared with that without AI assistance (p < 0.001; Table 3 and Figure 2).

**Figure 2.**
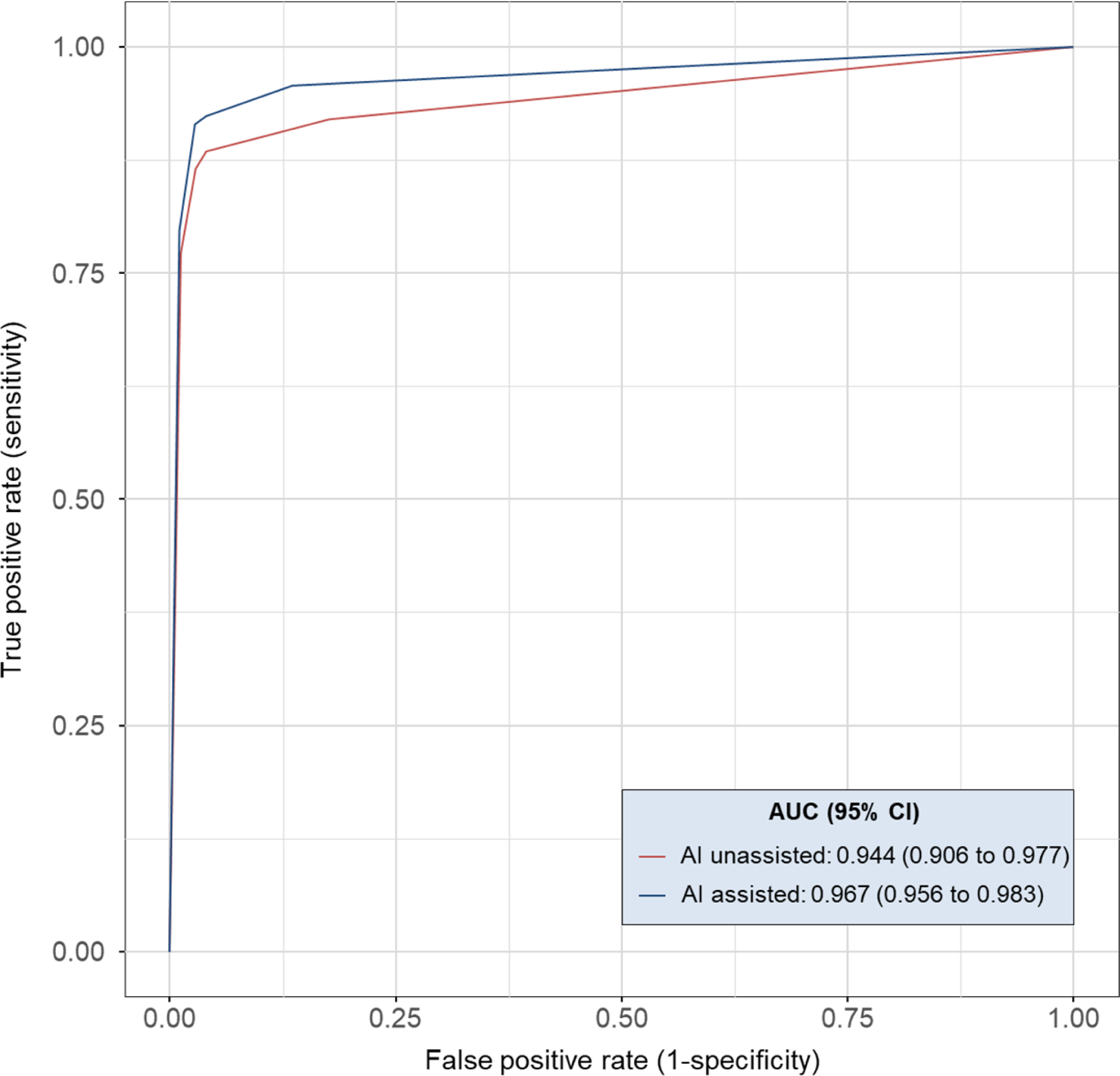
Diagnostic performance of readers without versus with artificial intelligence assist. Average reader receiver operating characteristic curves for detecting large vessel occlusion under two reading conditions: without and without AI assistance. Average area under the receiver operating characteristic curve (AUC) was computed across 4 readers participating in the study using the Obuchowski-Rockette, which accounts for the multireader multicase study design.

**Table 3.** Performance of readers without versus with artificial intelligence assist.

|  | With AI | Without AI | Difference | P |
| --- | --- | --- | --- | --- |
| Sensitivity |  |  |  |  |
| All reviewers | 91.82 (90.04 to 93.37) | 87.81 (85.73 to 89.68) | 4.00 (2.17 to 5.84) | < 0.001 |
| Reader 1 | 92.00 (88.14 to 94.92) | 80.00 (74.78 to 84.56) | 12.00 (7.43 to 16.57) | < 0.001 |
| Reader 2 | 86.55 (81.94 to 90.35) | 86.50 (81.87 to 90.31) | 0.00 (-3.50 to 3.50) | 1.00 |
| Reader 3 | 94.55 (91.16 to 96.92) | 92.00 (88.14 to 94.92) | 2.55 (-0.55 to 5.64) | 0.11 |
| Reader 4 | 94.18 (90.72 to 96.64) | 92.73 (88.99 to 95.50) | 1.45 (-1.73 to 4.64) | 0.37 |
| Specificity |  |  |  |  |
| All reviewers | 95.70 (94.44 to 96.74) | 96.30 (95.11 to 97.27) | -0.63 (-1.94 to 0.69) | 0.42 |
| Reader 1 | 96.56 (93.93 to 98.27) | 98.44 (96.39 to 99.49) | -1.87 (-4.16 to 0.41) | 0.11 |
| Reader 2 | 99.38 (97.76 to 99.92) | 98.13 (95.96 to 99.31) | 1.25 (-0.48 to 2.98) | 0.15 |
| Reader 3 | 93.08 (89.71 to 95.61) | 96.56 (93.93 to 98.27) | -3.46 (-6.26 to -0.66) | 0.016 |
| Reader 4 | 93.75 (90.51 to 96.14) | 92.26 (88.79 to 94.93) | 1.56 (-1.84 to 4.97) | 0.37 |
| Accuracy |  |  |  |  |
| All reviewers | 93.90 (92.86 to 94.83) | 92.36 (91.22 to 93.40) | 0.76 (0.01 to 1.50) | 0.049 |
| Reader 1 | 94.45 (92.30 to 96.15) | 89.92 (87.21 to 92.22) | 2.27 (0.69 to 3.85) | 0.007 |
| Reader 2 | 93.45 (91.15 to 95.30) | 92.76 (90.37 to 94.71) | 0.42 (-1.08 to 1.92) | 0.66 |
| Reader 3 | 93.76 (91.50 to 95.57) | 94.45 (92.30 to 96.15) | -0.34 (-1.72 to 1.04) | 0.72 |
| Reader 4 | 93.95 (91.72 to 95.73) | 92.48 (90.06 to 94.46) | 0.76 (-0.73 to 2.24) | 0.37 |

## Discussion

In this multicenter clinical trial, a fully automated AI software package for detecting LVO achieved a sensitivity of 86% and a specificity of 97%. For isolated MCA-M2 occlusion, the JLK-LVO software attained a sensitivity of 69% and a specificity of 96%. Additionally, the reader assessment study demonstrated that AI assistance significantly enhanced the sensitivity of LVO detection among residents in stroke care. To the best of our knowledge, this is the first clinical trial to prove the stand-alone efficacy and the improvement in reader performance using AI assistance across multicenter datasets in detecting LVO on CTA.

A few AI software packages have been implemented in clinical practice. RAPID LVO (iSchemaView, Menlo Park, CA), utilizing a traditional machine learning model that primarily relies on vessel density threshold assessment, showed higher performance in a pooled cohort from two stroke trials, with a sensitivity of 95% and specificity of 79%.^14^ However, RAPID LVO requires different threshold settings based on the occlusion site (ICA, M1, or M2), which may pose challenges for early-career physicians in interpreting the results. Furthermore, recent studies have demonstrated a lower PPV for RAPID LVO, which may contribute to alarm desensitization, leading to missed alarms or delayed responses.^15^ Viz LVO (Viz.AI, San Francisco, CA, USA) and CINA LVO (Avicenna.ai, La Ciotat, France), using an end-to-end deep learning algorithm to detect LVO, showed high sensitivity and specificity in recent clinical studies, with sensitivity ranging from 76% to 94%.^16–18^ In the present study, as a stand-alone tool, JLK-LVO achieved a sensitivity of 87% and a specificity of 97% in detecting intracranial LVO largely comparable to other software packages.

Recent studies have begun to shed light on the potential benefits of EVT for patients with MCA-M2 segment occlusions, expanding the traditional focus from proximal LVO to include more distal vessels. A meta-analysis revealed that patients with MCA-M2 occlusions treated with EVT showed improved functional outcomes at 90 days compared to those receiving standard medical therapy alone.^19^ Nevertheless, detecting isolated MCA-M2 occlusions remains challenging, even for experienced clinicians. In a study involving 520 patients with ischemic stroke, of which 16% had LVO and 40 patients had isolated MCA-M2 occlusion, experienced neuroradiologists missed 35% of MCA-M2 occlusions at initial CTA evaluation.^20^ Additionally, the study highlighted that non-neuroradiologists had a five-fold higher risk of missing LVO in a multivariable analysis.^20^ In this trial, JLK-LVO achieved a sensitivity of 69% in detecting isolated MCA-M2 occlusion, which is higher than that of other automated LVO detection software packages in patients with isolated MCA- M2 occlusion.^14–18^ This disparity may have resulted from the inclusion of isolated MCA-M2 occlusion as LVO in the training dataset for JLK-LVO.

The reader assessment study component of our research highlights another critical aspect of AI in stroke care: the improvement of diagnostic capabilities across varying levels of clinical experience. The observed improvement in sensitivity among early-career physicians when using AI assistance not only validates AI’s role as a diagnostic aid but also addresses a significant challenge in stroke care — the scarcity of vascular experts, especially in areas with limited resources. By offering a high degree of sensitivity and specificity in LVO detection, JLK-LVO can democratize access to high-quality stroke diagnostics, ensuring more patients are correctly identified for EVT, regardless of their geographical location or the immediate availability of stroke specialists.

Our study is subject to limitations inherent in its retrospective design and the potential for selection bias. Additionally, the real-world efficacy of the JLK-LVO software may vary due to differences in imaging equipment, protocols, and patient demographics across healthcare settings. Furthermore, the prevalence of LVO associated with intracranial arterial stenosis is notably higher in Asian populations,^21^ potentially introducing complexity into our trial dataset for deep learning analysis. This complexity arises because LVOs associated with intracranial arterial stenosis typically exhibit an increased number of collateral vessels^22^ and present with less distinct LVO characteristics when contrasted with LVOs resulting from cardioembolic occlusions. These factors underscore the need for prospective, multicenter studies to further validate our findings and explore the integration of JLK-LVO into diverse clinical workflows.

## Conclusion

In conclusion, we proved the clinical efficacy of JLK-LVO for detecting LVO in this multicenter clinical trial in patients with acute ischemic stroke. Furthermore, we demonstrated that AI-assisted reading significantly increases sensitivity of LVO detection in early-career physicians. By facilitating the early identification of patients eligible for EVT, JLK-LVO has the potential role to improve clinical outcomes of stroke patients.

## Data Availability

All data produced in the present study are available upon reasonable request to the authors

