## Supplementary Material for "Automated detection of large vessel occlusion using deep learning: a pivotal multicenter clinical trial and reader assessment study"

Jae Guk Kim et al.

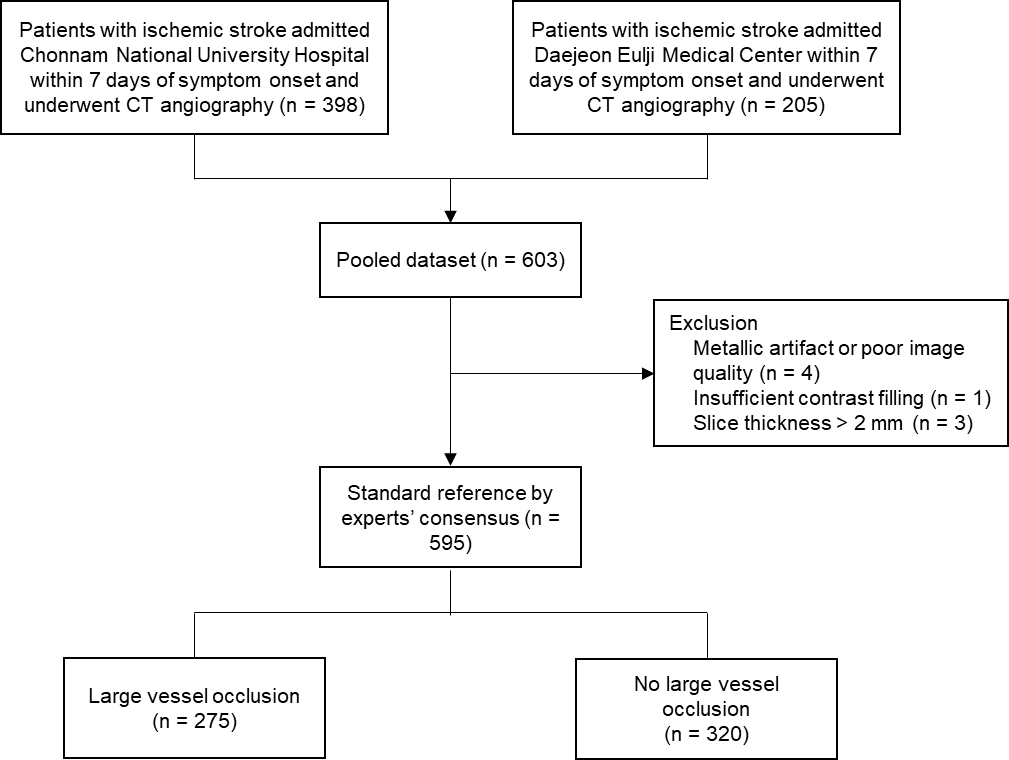

**Supplementary Figure 1. Study flow chart**

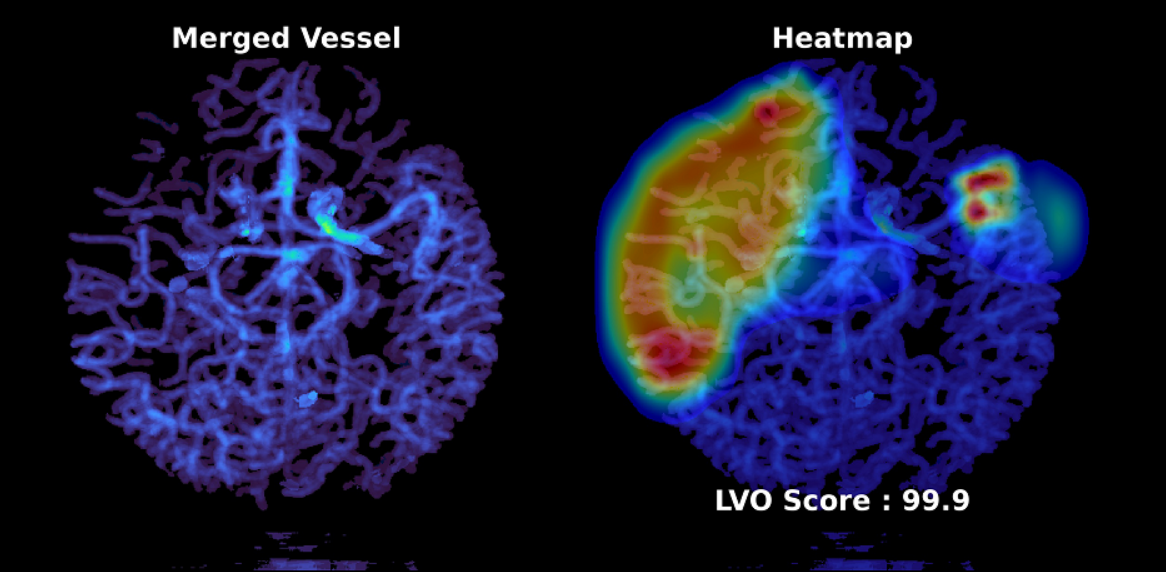

**Supplementary Figure 2. Merged vessel image, heatmap, and LVO probability score predicted by JBS-LVO**

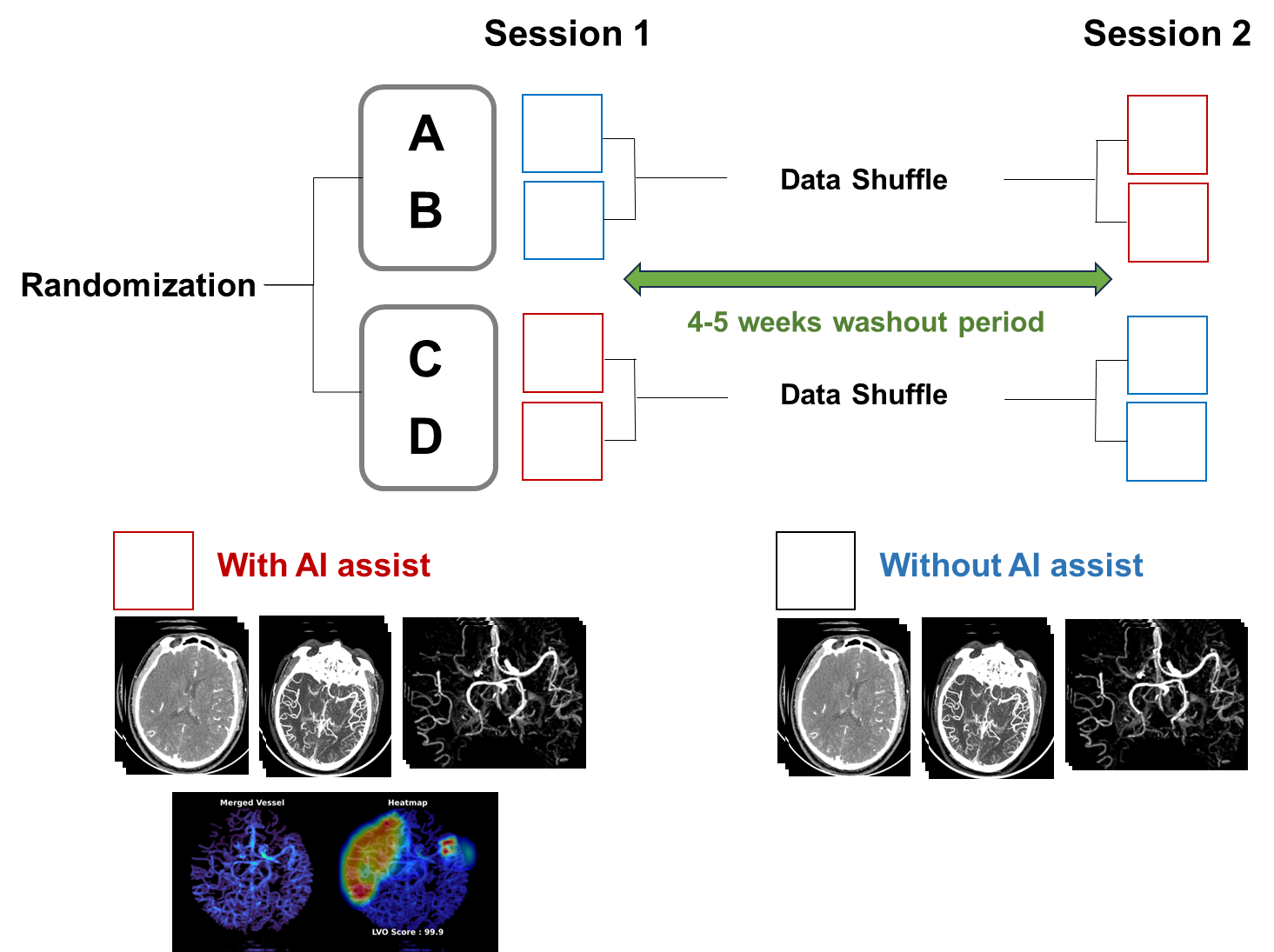

**Supplementary Figure 3. Schematic diagram showing reader assessment study.**

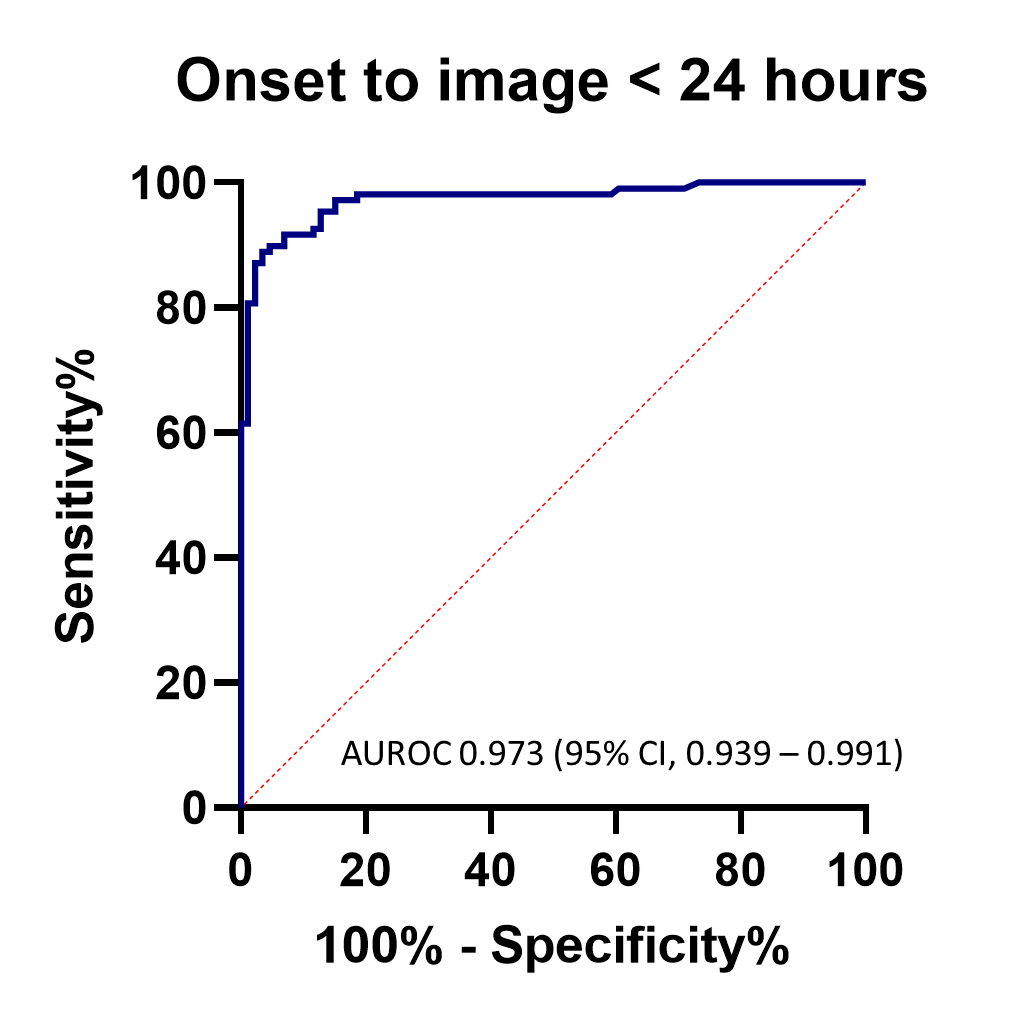

**Supplementary Figure 4. Area under the curve in patients with onset to image ≤ 24 hours.**

**Supplementary Table 1. Details of occlusion site**

| Occlusion site |  |
| --- | --- |
| Isolated intracranial ICA | 10 (3.6%) |
| Isolated MCA-M1 | 23 (8.4%) |
| Isolated MCA-M2 | 78 (28.4%) |
| MCA-M1 and MCA-M2 | 102 (37.1%) |
| Intracranial ICA and MCA-M1 | 3 (1.1%) |
| Intracranial ICA and MCA-M2 | 7 (2.5%) |
| Intracranial ICA, MCA-M1, and MCA-M2 | 46 (16.7%) |
| Bilateral | 6 (2.2%) |

ICA=internal carotid artery; MCA=middle cerebral artery

**Supplementary Table 2. Baseline characteristics stratified by participating centers**

|  | **CNUH (n = 395)** | **DEU (n = 200)** | **P value** |
| --- | --- | --- | --- |
| Age | 69.6 ± 13.6 | 66.7 ± 12.8 | 0.01 |
| Sex | 213 (53.9%) | 119 (59.5%) | 0.20 |
| Onset to admission, hr | 5.3 (2.1 – 21.3) | 2.8 (0.9 – 11.2) | < 0.001^a^ |
| Onset to imaging, hr | 54.3 (45.2 – 73.2) | 6.0 (1.9 – 17.5) | < 0.001^a^ |
| Initial NIHSS | 4 (1 – 10) | 4 (2 – 14) | < 0.001^a^ |
| Previous stroke | 65 (16.5%) | 36 (18.4%) | 0.56 |
| Hypertension | 209 (52.9%) | 130 (66.3%) | 0.002 |
| Diabetes | 100 (25.3%) | 68 (34.7%) | 0.017 |
| Atrial fibrillation | 112 (28.4%) | 52 (26.5%) | 0.64 |
| Occlusion site |  |  |  |
| Intracranial ICA | 31 (7.9%) | 34 (17.0%) | 0.001 |
| MCA-M1 | 101 (25.6%) | 73 (36.5%) | 0.006 |
| MCA-M2 | 146 (37.0%) | 87 (43.5%) | 0.12 |
| Isolated MCA-M2 occlusion^a^ | 53 (13.4%) | 9 (4.5%) | < 0.001 |
| Revascularization therapy | 71 (18.0%) | 89 (44.5%) | < 0.001 |
| Intravenous only | 41 (10.4%) | 8 (4.1%) |  |
| Endovascular only | 17 (4.3%) | 34 (17.4%) |  |
| Combined | 13 (3.3%) | 47 (24.0%) |  |

Data were presented as mean±SD, median (interquartile range), and number (percentage).

^a^Rank-sum test was used.

NIHSS=National Institute of Health Stroke Scale; ICA=internal carotid artery; MCA=middle cerebral artery.

**Supplementary Table 3. Performance of software after stratified by occlusion site**

|  | **Intracranial LVO** | | **Isolated MCA-M2 occlusion** | |
| --- | --- | --- | --- | --- |
| Confusion matrix | Prediction | | Prediction | |
|  | LVO | No LVO | LVO | No LVO |
| GT, LVO | 186 | 27 | 54 | 24 |
| GT, no LVO | 10 | 310 | 13 | 311 |
| Sensitivity (95% CI) | 0.873 (0.821 – 0.915) | | 0.692 (0.578 – 0.792) | |
| Specificity (95% CI) | 0.969 (0.943 – 0.985) | | 0.960 (0.932 – 0.978) | |
| PPV (95% CI) | 0.949 (0.908 – 0.975) | | 0.806 (0.691 – 0.892) | |
| NPV (95% CI) | 0.920 (0.886 – 0.940) | | 0.928 (0.895 – 0.954) | |

The parameters were estimated at the cutoff point of 0.5. LVO=large vessel occlusion; MCA=middle cerebral artery; GT=ground truth; PPV=positive predictive value; NPV=negative predictive value.
